# Keeping it close: The role of a Campus COVID Support Team (CCST) in sustaining a safe and healthy university campus during COVID-19

**DOI:** 10.1101/2022.05.01.22274540

**Authors:** Karen A McDonnell, Amita Vyas, Amanda Castel, Nitasha Nagaraj, Megan Landry

## Abstract

It has been over 24 months since the start of the COVID-19 pandemic forced university campuses to shut down and then reopen under new safety guidelines. Now as we move into the subsequent years of the pandemic, we can look back and evaluate what has worked, improvements to be made, and plans for providing a sustained response for a campus community. In this article we detail one campus response to the COVID-19 pandemic and directions being taken to ensure a sustained campus COVID support team (CCST) is in hand to ensure the health and safety of the university community. The CCST was created to serve as a one-stop-shop to help the university community navigate COVID-19 policies and procedures. The responsibilities of the CCST include conducting case investigations for any positive COVID-19 tests within the university community, contact tracing for authorized university affiliates, epidemiological surveillance and mitigation efforts, and communication through real-time analysis and dashboards. Continuous monitoring procedures demonstrated the CCST conducted all case investigations within the post-testing 24-hour window, thus keeping the university test-positivity rate below 3%. Quality improvement surveys demonstrated a high level of satisfaction with the CCST efforts and provided areas for improvement and sustainability. Having a public health faculty led CCST enabled the university to act swiftly when COVID-19 positive cases were emerging and deter widespread campus COVID-19 outbreaks. The CCST timeliness and connectivity to the campus has demonstrated benefits to the health and safety of the campus.

**Highlights:**

1. Universities are their own communities and having on campus COVID support teams can mitigate potential COVID-19 outbreaks.
2. Having a public health driven Campus COVID Support Team that can conduct case investigations within 24 hours of a positive test result has demonstrated benefits to taking responsive measures.
3. Continuous quality improvement efforts including surveys of the Campus COVID Support Team should be implemented for any COVID service efforts.

## Introduction

It has been over two years since the start of the COVID-19 pandemic forced university campuses to shut down and then reopen under new safety guidelines. As we move into subsequent years of the pandemic, it’s imperative to reflect on what has worked, improvements to be made, and plans for providing a sustained response for a campus community. In this article we detail one campus’ response to the COVID-19 pandemic and directions being taken to ensure sustained health and safety of the university community.

When the World Health Organization (WHO) declared COVID-19 a pandemic^1^ and the US went into a national shutdown, universities were forced to close their residential buildings and move to a virtual online format. As the spring semester served as a shock to the academic system, the summer months were replete with planning and strategy meetings to determine the forecast for campus engagement resuming in the fall of 2020. As the summer months of 2020 moved forward, the COVID-19 forecast became more concerning and few university campuses made the decision to open fully for the 2020-2021 academic year. Those that did open soon found themselves making difficult decisions when COVID-19 infections were emerging among students and staff. Now a year later, and with vaccines available to the college populations, the college community must sustain its enhanced safety efforts that were developed during the past year. As variants of the virus move through our populations, the protective measures that were initially developed in the midst of a crisis, are ones that should be sustained as demonstrated to be efficacious in the care of the university community.

## Background

Our own experience is with a metropolitan middle sized Mid-Atlantic university that has two main campuses, including a school of public health (SPH). When the pandemic was in the first wave for the geographic area in the spring of 2020, the SPH built upon their existing network with local health departments (LDOH) and met regularly to update the university administration on the real-time community efforts to mitigate the COVID-19 pandemic. These experiences were invaluable to the university planning efforts and provided informed options to proceed with campus reopening.

SPH faculty provided expertise in epidemiology and project management/evaluation to work with the LDOH leadership to establish initial contact tracing and case investigation task forces in each jurisdiction and to assist with epidemiological tracking of the novel coronavirus. The SPH faculty leaders of these efforts collectively came together to form the Campus COVID Support Team (CCST) to formulate the university response efforts. The CCST served as an integral part of the university response alongside safety and security, student services, athletics, student health, occupational health, and the newly designed Public Health Laboratory (PHL) COVID-19 PCR testing facility^2^. Upon reopening, the university mandated participation in regular surveillance COVID-19 testing and symptom checking surveys for the university community who have access to campus^2^.

The CCST was created to develop and implement university COVID-19 policies and to serve as a “one-stop-shop” to help the university community (students, staff, and faculty) navigate COVID-19 policies and procedures. The responsibilities of the CCST include:

- Conduct case investigations for any positive COVID-19 tests within the university community
- Conduct contact tracing for any positive COVID-19 tests for authorized university affiliates
- Monitoring and evaluation of COVID-19 testing protocols on the campus
- Advising on COVID-19 testing and quarantine/isolation policies/protocols
- Develop university communications and disseminate university metrics via an online dashboard

### Case Investigation

An important aspect of the CCST is to conduct a detailed case investigation and contact tracing protocol once individuals are confirmed as COVID-19 positive. A COVID-19 positive individual can be confirmed directly through regular or symptomatic testing at the PHL or can upload their PCR test result to a secure online portal. Within 24 hours of a confirmed positive test, a member of the CCST will call the individual and conduct an in-depth case investigation. At the time of contact, the individual is assured of their confidentiality and all data are entered into a secure RedCAP data portal. The case investigation procedures elicit information to assist the individual and inform the actions to be taken to restrict transmission of the virus. Figure 1 presents the categories of information gathered within the case-investigation. The CCST engages all necessary and identified resources to assist the positive individual and will set up a follow up schedule during the 10-day isolation period.

**Figure 1.**
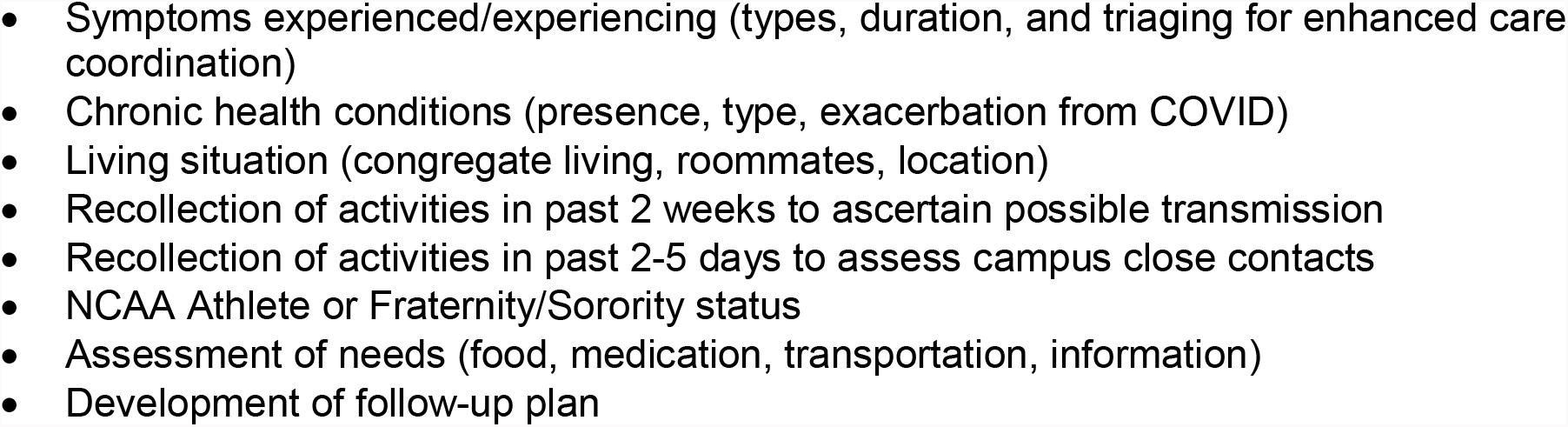
Campus Case Investigation Format.

### Campus Contact Tracing

Once the case investigation has been completed, trained Interview Assistants (IA) are provided with a list of university affiliated close contacts to interview. The CCST was permitted by the LDOH to conduct contact tracing only with university based close contacts. Close contacts are notified within 24 hours of the completed case investigation of their possible exposure and are instructed to monitor their symptoms, obtain a PCR COVID-19 test at the PHL in 3 to 5 days (or earlier if experiencing symptoms), and provided university and COVID-19 informational resources.

### Epidemiological Investigation

The CCST reviews all case data in real time, using epidemiological tools to conduct an outbreak investigation, and setting forth university recommendations to curb viral transmission. The CCST investigation and communication procedures in conjunction with serial surveillance through regular testing cadence and an onsite PCR testing facility, has provided the university with the tools to maintain a daily testing positivity rate below 3% and avoided shutdowns that have plagued other similar sized universities (see Figure 2 below). The testing cadence was initially set at a weekly cadence for all university personnel and reduced to biweekly once the semester demonstrated stability in a low-positivity testing outcome. The only exception was for athletes who followed the NCAA established testing guidelines.

**Figure 2:**
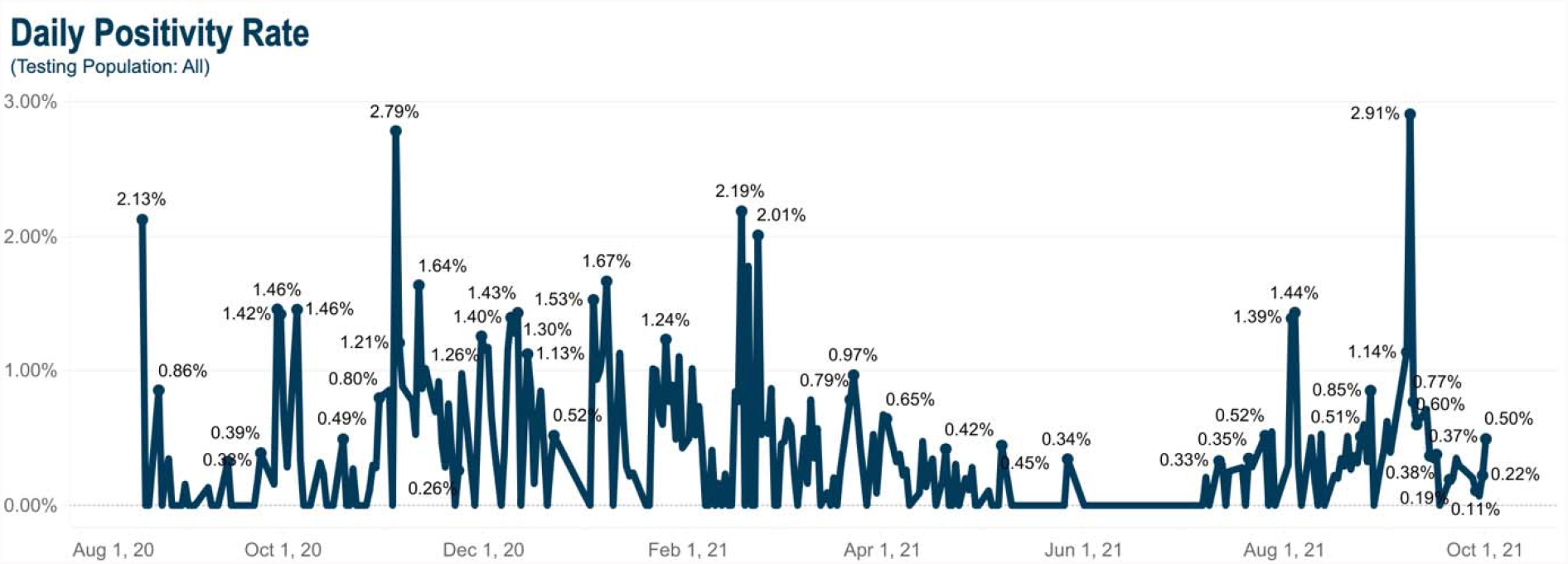
Daily Positivity Rate for All University Personnel from August 2020 to October 2021

### COVID Dashboard

Results of the real-time monitoring are communicated through the university COVID-19 Testing Dashboard (https://coronavirus.gwu.edu/). This dashboard is an interactive public site and provides aggregate and daily testing numbers, results, and positivity rates for the approximately 25,252 members of the on-campus community, who are required to take part in the regular testing protocol as a condition of access to campus. Currently, 5,272 faculty, staff and contractors and 19,980 students, including 6,500 residential students, are part of our campus community for the 2021-2022 academic year. The university has a COVID-19 vaccine mandate (allowing for medical and religious exemptions) and 98% of the university community is fully vaccinated.

### Quality Improvement

Our continuous monitoring procedures determined that the CCST conducted all case investigations within the 24-hour window after being notified by the PHL lab and medical authorizer (student/occupational health officer). As can be seen from Figure 2, the spikes in daily positivity for COVID-19 coincided with notable dates including back to campus move-in (August) and gatherings for holiday festivities (Labor Day, Thanksgiving break, Winter break, Spring break). As part of the prevention and mitigation efforts, the CCST was preemptive and sent out university communications reinforcing behavioral viral mitigation measures prior to holiday breaks in which students and staff have typically left the campus and engaged in larger social gatherings.

Initially, based on projections, the staffing of the CCST provided three Program Managers. However, as the census of the approved university cohort increased, so too did the need to add additional staff. With the full campus community, a total of 5 PMs were included on the team with a rotational schedule to assure at least one PM and one of the four faculty advisors are available to conduct case investigations as soon as the results are released (10am, 2pm, 7pm).

In addition, the CCST conducted a mid-year survey to obtain feedback on the CCST services and used these findings to engage in quality and process improvements. In December 2020, CCST designed the “CCST Experience Survey”, to determine satisfaction with the CCST provided services. To participate in the survey, individuals had to be university community members who had tested positive for COVID-19 and participated in a case investigation with the CCST. Individuals completed an online 5-10 minute survey, no identifying information was collected to maintain anonymity via a secured link sent via their campus email address.

Respondents were asked to provide their university affiliation (student, staff, faculty, contractor), their age range, gender (male, female, non-binary), and answer twelve questions about the services provided by the CCST. The first set of 10 questions asked about service quality and level of agreement for each item was assessed on a 5-point Likert scale (5=strongly agree to 1=strongly disagree). Two questions were asked to assess the overall level of satisfaction with services provided (5-point Likert scale with 5= extremely satisfied and 1= extremely dissatisfied) and a second question about the overall experience with the CCST (5-point Likert scale 5= excellent to 1= terrible). In addition, an open text box was provided for additional comments and suggestions. Lastly, as the CCST interaction was in addition to the LDOH case investigation, the survey asked how long after testing positive for COVID-19 did the interaction with LDOH occur.

## Results

A total of 75 individuals completed the survey, with the majority of the sample identifying as a student (61%), female (56%), and between the ages of 18-24 (58%) (see Table 1). The majority of the respondents (n=63) were contacted by the LDOH with an overage contact time of 4.02 days (range same day to 23 days).

**Table 1:** Characteristics of CCST Quality Improvement Survey. (n=75)

| Role at the University | N(%) |
| --- | --- |
| Faculty | 6 (8.0) |
| Staff/Contractor | 23 (30.7) |
| Student | 46 (61.3) |
| **Gender** |  |
| Male | 33 (44.0) |
| Female | 42 (56.0) |
| **Age** |  |
| 18-24 years | 44 (58.7) |
| 25-34 years | 8 (10.7) |
| 35-44 years | 9 (12.0) |
| 45-54 years | 5 (6.7) |
| 55 and older | 9 (12.0) |

Table 2 presents the mean responses for each of the 10 service quality items and two satisfaction items. The mean response for the service quality items ranged from a low of 4.05 (sd=1.02) when asked about feeling less overwhelmed and less stressed after the CCST call to a high of 4.81 for two items asking if the CCST staff treated the respondent with respect and the CCST staff provided clear information as to why the person was being contacted. When asked about the level of satisfaction with the services provided by the CCST, the mean response was 4.70 and the mean for satisfaction with the overall CCST experience was 4.63. Reported levels of quality improvement and satisfaction for all items did not differ by respondent university affiliation, gender, or age.

**Table 2.**
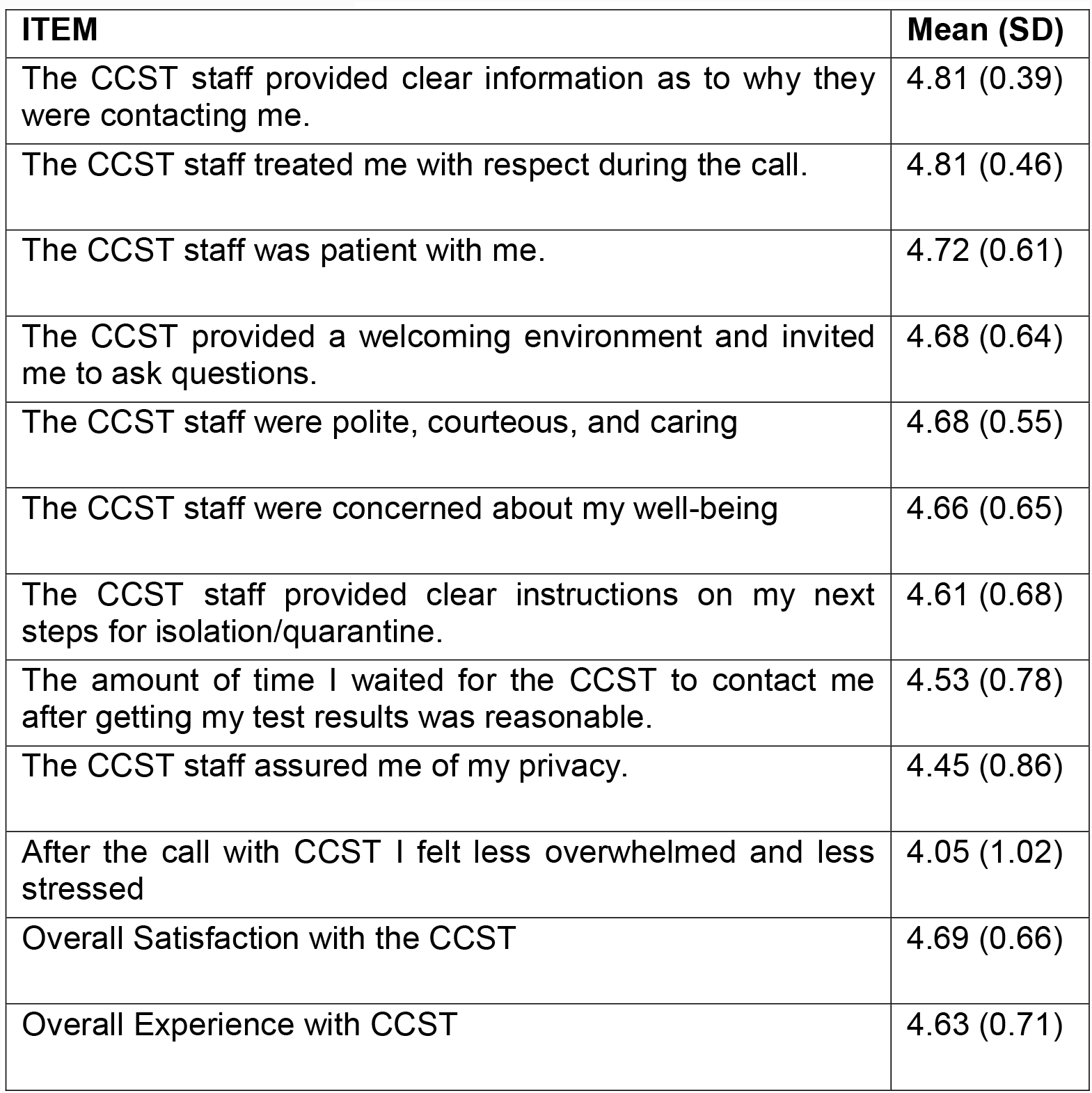
CCST Quality Improvement Items. *Thinking back on your experience with the CCST, please rate your level of agreement with the following statements:*

| **ITEM** | **Mean (SD)** |
| --- | --- |
| The CCST staff provided clear information as to why they were contacting me. | 4.81 (0.39) |
| The CCST staff treated me with respect during the call. | 4.81 (0.46) |
| The CCST staff was patient with me. | 4.72 (0.61) |
| The CCST provided a welcoming environment and invited me to ask questions. | 4.68 (0.64) |
| The CCST staff were polite, courteous, and caring | 4.68 (0.55) |
| The CCST staff were concerned about my well-being | 4.66 (0.65) |
| The CCST staff provided clear instructions on my next steps for isolation/quarantine. | 4.61 (0.68) |
| The amount of time I waited for the CCST to contact me after getting my test results was reasonable. | 4.53 (0.78) |
| The CCST staff assured me of my privacy. | 4.45 (0.86) |
| After the call with CCST I felt less overwhelmed and less stressed | 4.05 (1.02) |
| Overall Satisfaction with the CCST | 4.69 (0.66) |
| Overall Experience with CCST | 4.63 (0.71) |

Nineteen (24%) of the respondents provided qualitative recommendations. Overall, the qualitative responses were positive in content and supported the current processes of the CCST. Qualitative remarks did provide insight for improvements including the number and frequency of calls received by the CCST. In general, these comments pertained to a combination of calls from the CCST and LDOH conducting their case investigation. While the CCST conducted a timely case investigation within 24 hours of the notification of a positive COVID-19 test result, respondents noted that if contacted by the LDOH, this contact was delayed an average of 4 days. Respondents noted that the number of calls were overwhelming, especially if experiencing negative symptoms.

These comments were helpful to the CCST and modifications were made to the check-in calls with the number of calls for those not experiencing emerging illness dropping from an average of 2 calls at 5 and 10 days after the initial case interview, to conducting one, brief check-in call around day 7. The CCST modified the exit portion of the case investigation to manage communication expectations and outlined who should be calling, when, and for what purpose.

## Conclusion

Having a functioning CCST led by public health faculty enabled the university to act swiftly when COVID-19 positive cases were emerging. The CCST monitored the on-campus testing status and when more than one positive test was found from the PHL, could utilize the case investigation and contact tracing social mapping tools to monitor potential instances of COVID transmission, with a particular emphasis for campus situations and off-campus events. Through our timely case investigations and campus contact tracing, the CCST was able to notify and engage positive cases in their care coordination and provide their close contacts with timely notifications and ability to obtain on demand COVID-19 testing. The capability to contact persons testing positive within 24 hours and immediately start the contact tracing process has demonstrated benefits to the health and safety of the campus that would not have been realized had the CCST not been an active participant in the process.

## Data Availability

All data produced in the present work are contained in the manuscript and available online at coronavirus.gwu.edu

https://coronavirus.gwu.edu/

## Abbreviations

